# Severity of Prior COVID-19 Infection is Associated with Postoperative Outcomes Following Major Inpatient Surgery

**DOI:** 10.1101/2023.04.12.23288412

**Authors:** Nathaniel B. Verhagen, Gopika SenthilKumar, Taylor Jaraczewski, Nicolas K. Koerber, Jennifer R. Merrill, Madelyn A. Flitcroft, Aniko Szabo, Anjishnu Banerjee, Xin Yang, Bradley W. Taylor, Carlos E. Figueroa Castro, Tina W.F. Yen, Callisia N. Clarke, Kathryn Lauer, Kurt J. Pfeifer, Jon C. Gould, Anai N. Kothari, N3C Consortium

**Author notes:** **Corresponding Author:** Anai N. Kothari, MD MS FSSO Medical College of Wisconsin Division of Surgical Oncology 8701 W Watertown Plank Road Milwaukee, Wisconsin 53226, USA (414)955-1443.

## Abstract

**Objective:** To determine the association between severity of prior history of SARS-CoV-2 infection and postoperative outcomes following major elective inpatient surgery.

**Summary Background Data:** Surgical guidelines instituted early in the COVID-19 pandemic recommended delay in surgery up to 8 weeks following an acute SARS-CoV-2 infection. Given that surgical delay can lead to worse medical outcomes, it is unclear if continuation of such stringent policies is necessary and beneficial for all patients, especially those recovering from asymptomatic or mildly symptomatic COVID-19.

**Methods:** Utilizing the National Covid Cohort Collaborative (N3C), we assessed postoperative outcomes for adults with and without a history of COVID-19 who underwent major elective inpatient surgery between January 2020 and February 2023. COVID-19 severity and time from SARS-CoV-2 infection to surgery were each used as independent variables in multivariable logistic regression models.

**Results:** This study included 387,030 patients, of which 37,354 (9.7%) had a diagnosis of preoperative COVID-19. History of COVID-19 was found to be an independent risk factor for adverse postoperative outcomes even after a 12-week delay for patients with moderate and severe SARS-CoV-2 infection. Patients with mild COVID-19 did not have an increased risk of adverse postoperative outcomes at any time point. Vaccination decreased the odds of mortality and other complications.

**Conclusions:** Impact of COVID-19 on postoperative outcomes is dependent on severity of illness, with only moderate and severe disease leading to higher risk of adverse outcomes. Existing wait time policies should be updated to include consideration of COVID-19 disease severity and vaccination status.

## INTRODUCTION

As of February 2023, over 100 million cases of SARS-CoV-2 infections have been reported in the US^1^. Among those with prior infection, 15–20% will require elective surgery^2^. Prior history of COVID-19 infection is associated with adverse postoperative outcomes and complications^3–6^, and this risk can persist up to eight weeks after infection^5^. Accordingly, early in the pandemic, several groups recommended a delay in surgery for 7–8 weeks following a COVID-19 infection^5, 7^.

While these guidelines attempted to mitigate the risk of COVID-19 by delaying surgery, as the pandemic transitions to an endemic phase, there is a paucity of data to guide the de-implementation of these policies. This is especially important when also considering that delaying care can worsen both short- and long-term health outcomes^8^. In addition, delayed surgical care can lead to prolonged poor quality of life including chronic pain, impaired mobility, and inability to work.

The severity of a SARS-CoV-2 infection can vary widely from mild flu-like symptoms or no symptoms to systemic disease requiring hospitalization and even critical care. Most current guidelines for managing COVID-19 in the perioperative setting are not stratified by disease severity; however, there appears to be a relationship between severity of infection and postoperative outcomes^8^. Prior studies also show that compared to those with asymptomatic or mild disease, postoperative intensive care unit stay is higher among those with severe COVID-19 who require hospitalization^9^. This suggests that including severity of SARS-CoV-2 infection in surgical wait time guidelines may facilitate more personalized patient recommendations and prevent unnecessary delays to surgery for those with less severe infections. In addition, since disease severity can be lowered through vaccination^2^, and as of February 2023, over 269 million people in the US received at least 1 dose of COVID-19 vaccine^1^, the impact of preoperative vaccination among patients who present with COVID-19 prior to requiring surgery must be taken into consideration.

The primary objective of this study was to measure the association between the severity of SARS-CoV-2 infection prior to major elective surgery and postoperative adverse outcomes. Secondary objectives included investigating how severity should be considered in surgical wait time guidelines and the influence of vaccination on postoperative outcomes.

## METHODS

Electronic health record data from the National COVID Cohort Collaborative (N3C), the largest multi-site centralized data source of COVID-19 patients in the United States, was retrospectively analyzed using the N3C Data Enclave. PCORnet, TriNetX, OMOP, and ACT are the primary data sources that contribute to N3C. A detailed description of the N3C’s rationale, design, infrastructure, and deployment has been previously reported. Patients 18 years and older who underwent major inpatient surgery between January 2020 and February 2023 were included, and the surgical procedures were selected based on prior literature assessing the role of COVID-19 on surgical outcomes^5^. The following were included: hip arthroplasty, knee arthroplasty, laminectomy, spinal arthrodesis, craniectomy, aortic aneurysm repair, lung excision, coronary artery bypass graft, esophagectomy, mastectomy, prostatectomy, colectomy, gastrectomy, hepatectomy, and pancreatectomy (Supplemental Table 1).

Patient characteristics (sex, age, race and ethnicity, Charlson Comorbidity Index)^11^, were identified using the N3C’s Shared Knowledge Store patient fact table. Standard SNOMED codes from the Observational Health Data Science and Informatics’ (OHDSI) ATLAS tool were used to identify elective inpatient procedures and patients’ comorbidities (Supplemental Table 1,2). Exclusion criteria included undergoing natural orifice, percutaneous, endoscopic, diagnostic (unless open surgery), or transplant procedures. For patients with multiple surgeries, the first surgery after SARS-CoV-2 infection was used.

COVID-19 positivity was defined as having a positive lab measurement (PCR or antigen) or a positive COVID-19 diagnosis (ICD10-CM code U07.1) any time before surgery. Patients with COVID-19 within 24 hours of surgery were excluded. The severity of a patient’s COVID-19 was determined using the World Health Organization’s (WHO) Clinical Progression Scale (CPS)^11^. Patient-specific severity was obtained from the N3C Knowledge Store and derived from critical visits associated with COVID positivity. Mild severity was defined as having outpatient visit only (WHO Severity 1–3). Moderate severity was defined as requiring hospitalization without pulmonary support (WHO Severity 4–6). Severe disease was defined as needing hospitalization with extracorporeal membrane oxygenation (ECMO or invasive ventilation, WHO Severity 7– 9)^12^. The N3C Knowledge Store was used to determine the vaccination status of patients. Fully vaccinated patients were defined as having either received a single dose of the viral vector vaccine or two doses of mRNA vaccine at least 14 days before their first SARS-CoV-2 infection. Patients who received only one dose of the mRNA vaccine or were vaccinated after a COVID-19 diagnosis were not considered fully vaccinated.

The primary outcome of the study was a 30-day composite adverse postoperative event and included the following: mortality, unplanned readmission, acute myocardial infarction, cardiac arrhythmia, deep vein thrombosis, pneumonia, pulmonary embolism, renal failure, respiratory failure, sepsis, and urinary tract infection. Each individual 30- day complication included in the composite outcome was also separately evaluated. Outcomes were constructed using SNOMED codes and concepts associated with chronic conditions or directly caused by COVID-19 were excluded (Supplemental Table 3).

Unadjusted baseline characteristics and comparisons between patients with and without prior COVID-19 were analyzed using standard descriptive statistics. Adjusted analyses were performed using multivariable logistic regression models. Separate models were fit for each outcome with covariates that included age, sex, race and ethnicity, Charlson Comorbidity Index (CCI), and relative surgical risk. Relative surgical risk is a computed feature based on the expected median length of stay for a given procedure. Additional logistic regression models were fit to measure the association between 30-day postoperative adverse outcomes and (1) time between SARS-CoV-2 infection and surgery, (2) disease severity, and (3) vaccination status. Within each disease severity group, the risk of 30-day postoperative adverse outcomes as a factor of time between infection and surgery was analyzed. All analysis were conducted using the N3C Data Enclave using the tidyverse, ggplot2, gtsummary, splines, and broom R packages.

## RESULTS

387,030 patients were included in the study and 37,354 (9.7%) had a preoperative diagnosis of COVID-19. Table 1 details baseline patient demographics and characteristics in those with and without a history of COVID-19.

**Table 1.** Patient characteristics and 30-day postoperative adverse outcomes in patients with and without preoperative COVID-19

| Characteristics | No Hx of COVID-19<br>(N = 349,676),<br>No. (%) | Hx of COVID-19<br>(N = 37,354),<br>No. (%) | P value |
| --- | --- | --- | --- |
| **Patient Characteristics** |  |  |  |
| Age, years |  |  |  |
| **Median, (Q1-Q3)** | 63 (52, 72) | 61 (49, 70) | <0.001 |
| **18-29** | 15166 (4.3%) | 1625 (4.4%) | <0.001 |
| **30-49** | 60828 (17%) | 7734 (21%) |  |
| **50-64** | 111160 (32%) | 12838 (34%) |  |
| **65+** | 162522 (46%) | 15157 (41%) |  |
| Sex |  |  | <0.001 |
| **Female** | 190052 (54%) | ‡20952 (≈56%) |  |
| **Male** | 159505 (46%) | ‡16399 (≈44%) |  |
| **Missing or Unknown** | 119 (<0.1%) | 20 or fewer |  |
| Race and Ethnicity |  |  | <0.001 |
| **Non-Hispanic White** | 257334 (74%) | 25833 (69%) |  |
| **Non-Hispanic Black** | 42740 (12%) | 4867 (13%) |  |
| **Non-Hispanic Asian** | 8395 (2.4%) | 810 (2.2%) |  |
| **Hispanic** | 21325 (6.1%) | 3625 (9.1%) |  |
| **Other or Unknown** | 19930 (5.7%) | 2403 (6.4%) |  |
| Smoking Status |  |  | <0.001 |
| **Nonsmoker** | 304141 (86%) | 33151 (89%) |  |
| **Current or Former Smoker** | 47716 (14%) | 4203 (11%) |  |
| Charlson Comorbidity Index (CCI) |  |  | <0.001 |
| **0** | 72432 (21%) | 6772 (18%) |  |
| **1** | 42341 (12%) | 4412 (12%) |  |
| **2** | 63704 (18%) | 6343 (17%) |  |
| **3** | 38900 (11%) | 4275 (11%) |  |
| **4+** | 132299 (38%) | 15552 (42%) |  |
| Comorbidities |  |  |  |
| **CKD** | 22374 (6.4%) | 3862 (10%) | <0.001 |
| **CHF** | 20816 (6.0%) | 3454 (9.2%) | <0.001 |
| **COPD** | 21089 (6.0%) | 3134 (8.4%) | <0.001 |
| **Coronary Artery Disease** | 39221 (11%) | 5852 (16%) | <0.001 |
| **Depression** | 35956 (10%) | 5875 (16%) | <0.001 |
| **Diabetes** | 41584 (12%) | 7144 (19%) | <0.001 |
| **GERD** | 58830 (17%) | 9432 (25%) | <0.001 |
| **Hypertension** | 100981 (29%) | 14669 (39%) | <0.001 |
| **Liver Disease** | 5730 (1.6%) | 977 (2.6%) | <0.001 |
| **Obesity** | 51446 (15%) | 8920 (24%) | <0.001 |
| Relative Risk of Surgery |  |  | 0.058 |
| **Low Risk** | 134385 (38%) | 14364 (38%) |  |
| **Medium Risk** | 166702 (48%) | 17642 (47%) |  |
| **High Risk** | 48589 (14%) | 5348 (14%) |  |
| Vaccine Status |  |  | <0.001 |
| **Fully Vaccinated** | 48441 (14%) | 5381 (14%) |  |
| **Non-Fully Vaccinated** | 301235 (86%) | 31973 (86%) |  |
| **30-day postoperative adverse outcomes** |  |  |  |
| Composite Adverse Event | 75075 (21.5%) | 8493 (22.7%) | <0.001 |
| Mortality | 4791 (1.4%) | 666 (1.8%) | <0.001 |
| Hospital Readmission | 38175 (11%) | 4248 (11%) | 0.008 |
| Any Non-Fatal Complication | 73555 (21%) | 8257 (22%) | <0.001 |
| Acute Myocardial Infarction | 9089 (2.6%) | 1020 (2.7%) | 0.13 |
| Cardiac Arrhythmia | 34326 (9.8%) | 3752 (10%) | 0.16 |
| Deep Vein Thrombosis | 4648 (1.3%) | 532 (1.4%) | 0.13 |
| Pneumonia | 9623 (2.8%) | 1093 (2.9%) | 0.053 |
| Pulmonary Embolism | 4296 (1.2%) | 519 (1.4%) | 0.008 |
| Renal Failure | 22488 (6.4%) | 2592 (6.9%) | <0.001 |
| Respiratory Failure | 18862 (5.4%) | 2017 (5.4%) | 0.97 |
| Sepsis | 10244 (2.9%) | 1188 (3.2%) | 0.007 |
| Urinary Tract Infection | 9662 (2.8%) | 1045 (2.8%) | 0.71 |

‡ To comply with N3C policy, counts below 20 are displayed as <20 and additional values were skewed by up to 5 in order to render it impossible to back-calculate precise counts in the 'Missing or Unknown' category. Hx: history; CKD: chronic kidney disease; CHF: congestive heart failure; COPD: chronic obstructive pulmonary disease; GERD: gastroesophageal reflux disease.

### History of COVID-19 and 30-day postoperative adverse outcomes

On unadjusted analysis, rate of 30-day postoperative adverse events was greater in patients with a history of COVID-19 than those without (22.7% vs. 21.5%, P<0.001, Table 1). Additionally, the rates of 30-day mortality (1.8% vs. 1.4%), postoperative pulmonary embolism (1.4% vs. 1.2%; P=0.008), renal failure (6.9% vs. 6.4%; P<0.001), sepsis (3.2% vs. 2.9%; P=0.007), and composite non-fatal event (22% vs. 21%; P<0.001) were all significantly greater in patients with a history of COVID-19 (Table 1). Following adjustment for age, sex, race, ethnicity, smoking status, comorbid disease, and relative risk of surgery, patients with a history of COVID-19 had greater odds of 30- day composite adverse postoperative events (aOR 1.09 [1.06–1.12]; Figure 1).

**Figure 1.**
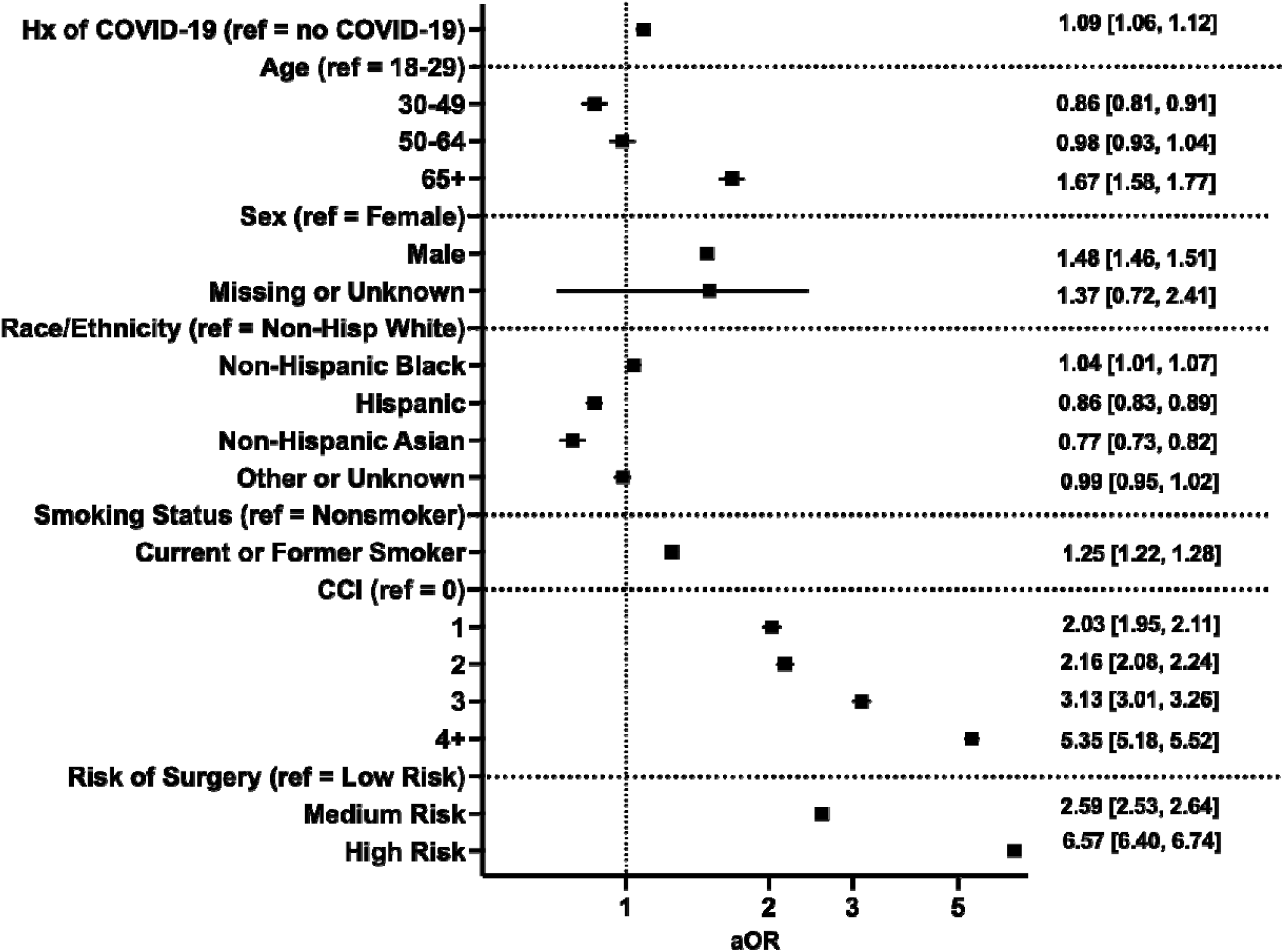
Multivariable regression assessing the association between prior history of COVID-19 and composite 30-day postoperative adverse event. Hx: history; CCI: Charlson Comorbidity Index; aOR: adjusted odds ratio (with 95% confidence interval).

### Association of time between SARS-CoV-2 infection and surgery and 30-day postoperative adverse outcomes

Patients were stratified based on timing of surgery relative to SARS-CoV-2 infection. Groups included 0–4 weeks (N=7,425, 19.9%), 4–8 weeks (N=3,936, 10.5%), 8–12 weeks (N=2,604, 7.0%) and 12+ weeks (N=23,389, 62.6%). Patients who underwent surgery 0–4 weeks after COVID-19 had overall higher rates of postoperative adverse events, including mortality (3.3% vs. 1.4%; P<0.001) and any non-fatal complications (26% vs. 21%, P<0.001), when compared to patients without COVID-19 (Supplemental Table 4). On adjusted analysis (Figure 2, Supplemental Table 5), patients who underwent surgery within 4 weeks of SARS-CoV-2 infection had increased odds of 30- day postoperative composite adverse event (aOR 1.27 [1.20–1.35]), mortality (aOR 2.34 [2.05–2.67]), deep vein thrombosis (aOR 1.33 [1.12–1.57]), pneumonia (aOR 1.60 [1.43–1.79]), pulmonary embolism (aOR 1.44 [1.20–1.70]), renal failure (aOR 1.28 [1.17–1.39]), respiratory failure (aOR 1.53 [1.40–1.66]), and sepsis (aOR 1.52 [1.36– 1.69]). The odds of composite adverse postoperative outcome returned to baseline at 12 weeks following infection (aOR 1.01 [0.98–1.05] at 12+ weeks).

**Figure 2.**
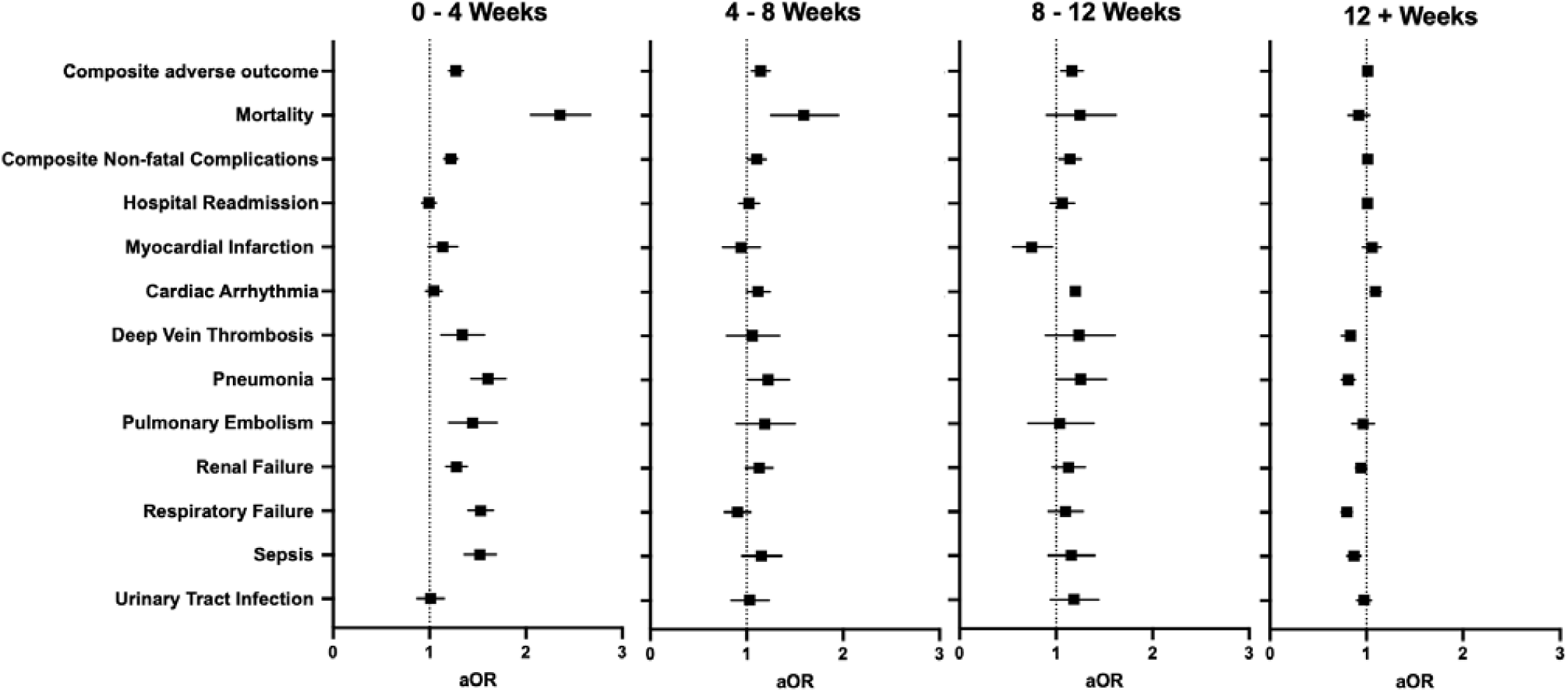
Multivariable regression assessing the association between timing of infection relative to surgery and 30-day postoperative adverse events. All models adjusted for age, sex, race and ethnicity, smoking status, comorbid disease, and relative risk of surgery. aOR: adjusted odds ratio (with 95% confidence interval).

### Association of severity of SARS-CoV-2 infection and 30-day postoperative adverse outcomes

Patients were next stratified based on the severity of their SARS-CoV-2 infection prior to surgery, and 31,677 (84.8%), 5,009 (13.4%), and 655 (1.8%) patients had mild, moderate, and severe disease, respectively. Incidence of adverse 30-day postoperative outcomes increased with severity across all measured outcomes (Supplemental Table 6), and patients with severe disease had the highest rate of mortality (14% vs. 1.4%), any non-fatal complication (60% vs. 21%), and hospital readmission (18% vs. 11%) when compared to patients without COVID-19 history (P<0.001 all). After adjusting for patient demographics, risk of surgery, and comorbidities, patients with moderate and severe disease had higher odds for all assessed adverse events except for pulmonary embolism for moderate COVID-19 (aOR 1.19 [0.96–1.45]) and hospital readmission for severe COVID-19 (aOR 1.20 [0.98–1.46]) (Figure 3, Supplemental Table 7).

**Figure 3.**
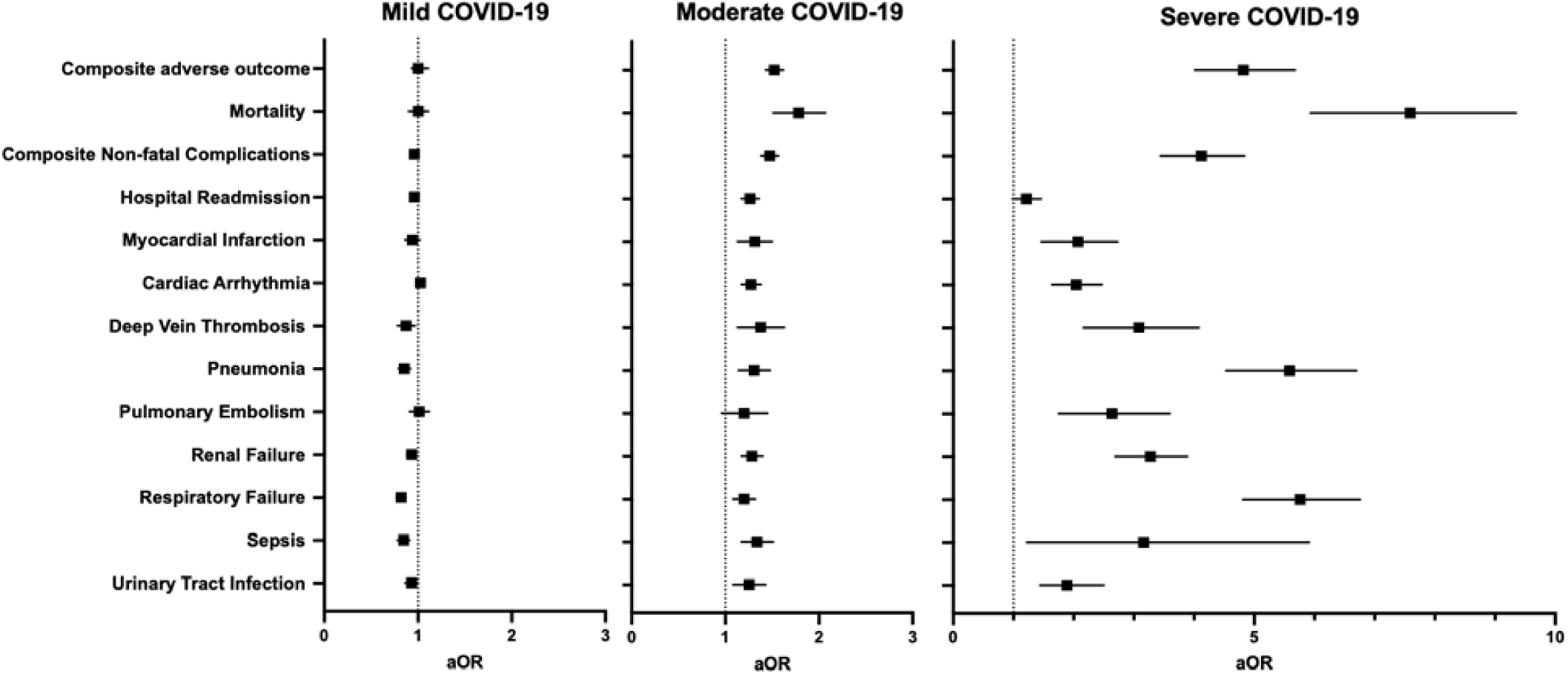
Multivariable regression assessing the association between COVID-19 disease severity and 30-day postoperative adverse outcomes. All models adjusted for age, sex, race and ethnicity, smoking status, comorbid disease, and relative risk of surgery. aOR: adjusted odds ratio (with 95% confidence interval).

### The interplay between timing of surgery and COVID-19 disease severity in influencing risk for 30-day composite postoperative complications

For each COVID-19 severity group (mild, moderate, severe), the adjusted odds of having a composite adverse postoperative event were measured (Figure 4). Patients with mild COVID-19 infections did not demonstrate an increased risk, regardless of the time between infection and surgery. Patients with moderate disease were observed to have increased risk for composite adverse events at 0–4 weeks (aOR 1.84 [1.61–2.10]) that persisted beyond 12 weeks between COVID-19 and surgery (aOR 1.36 [1.25– 1.48]) when compared to patients without prior history of COVID-19. The magnitude of risk was greater for patients with severe disease, with highest odds for an adverse event occurring if surgery was performed within 4 weeks of SARS-CoV-2 infection (aOR 8.41 [6.42–11.1]). This risk did decrease with time, though it remained elevated when compared to patients who did not have prior COVID-19 (aOR 2.22 [1.66–2.95] at 12+ weeks).

**Figure 4.**
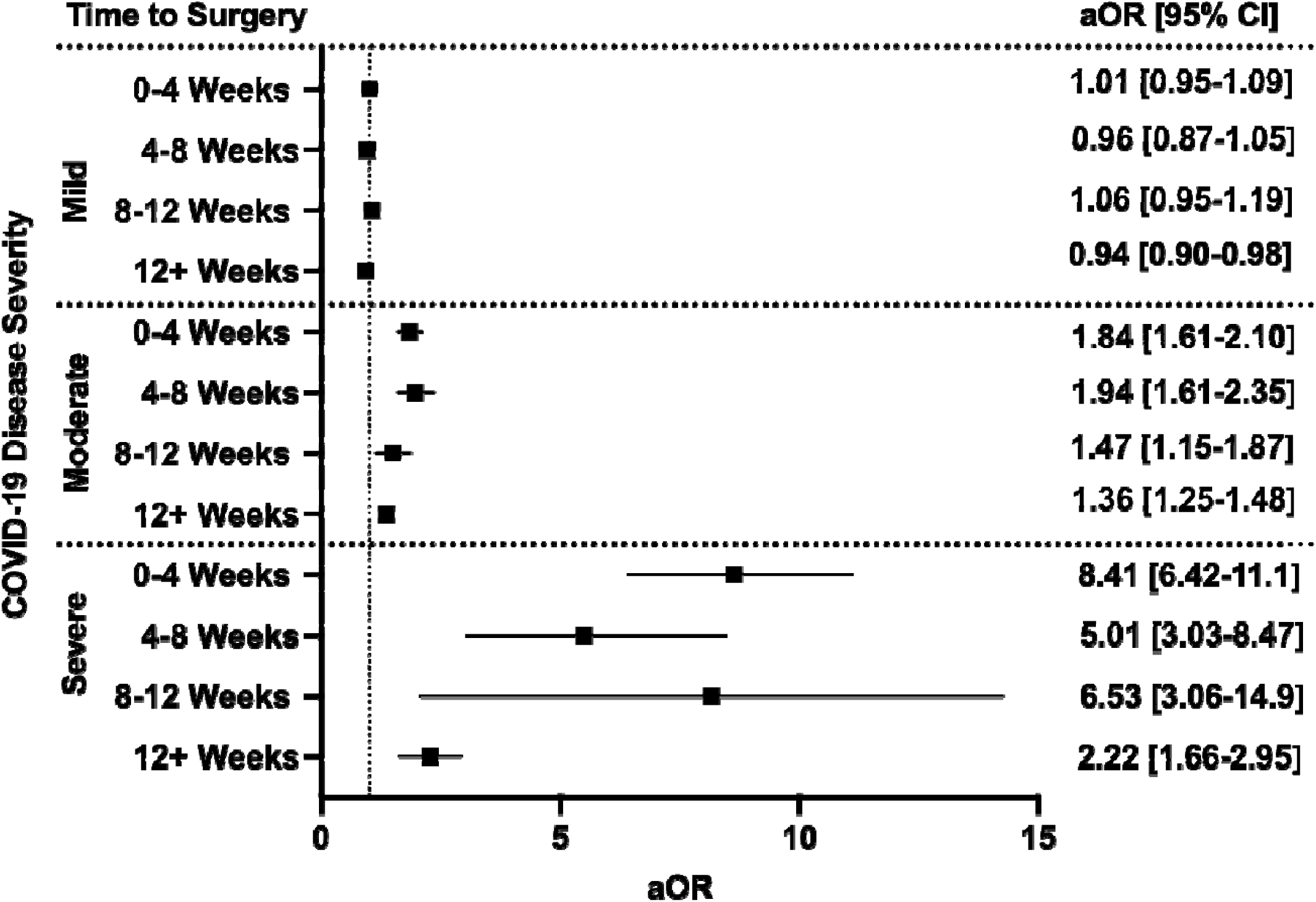
The interplay between timing of surgery and COVID-19 disease severity in influencing odds of composite 30-day postoperative adverse events. All models adjusted for age, sex, race and ethnicity, smoking status, Charlson Comorbidity Index, and relative risk of surgery. aOR: adjusted odds ratio (with 95% confidence interval).

### Preoperative vaccination and 30-day postoperative adverse outcomes

In patients with COVID-19 prior to surgery, vaccination significantly decreased rates of pneumonia (2.3% vs. 3.0%; P=0.003), respiratory failure (4.2% vs. 5.6%; P<0.001), and sepsis (2.7% vs. 3.3%; P<0.05) (Supplemental Table 8). After adjustment for patient demographics, comorbidities, and relative risk of surgery, vaccination was associated with decreased odds of mortality (aOR 0.76 [0.59–0.95]), pneumonia (aOR 0.73 [0.60– 0.88]), respiratory failure (aOR 0.72 [0.62–0.83]), and sepsis (aOR 0.79 [0.66–0.94]) (Figure 5).

**Figure 5.**
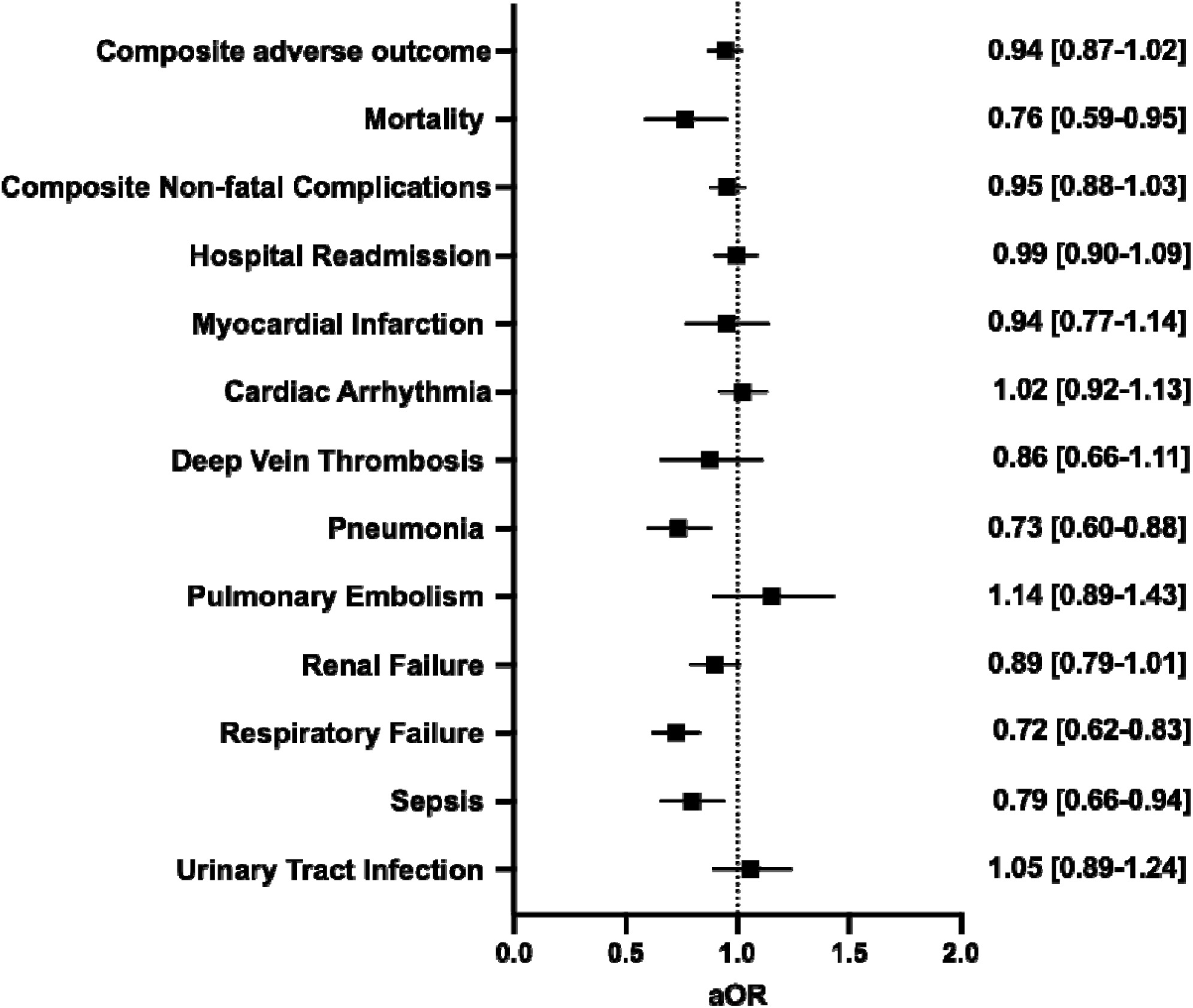
Multivariable regression assessing the association between full vaccination in patients with a history of COVID-19 and 30-day postoperative adverse events. All models adjusted for age, sex, race and ethnicity, smoking status, Charlson Comorbidity Index, and relative risk of surgery. aOR: adjusted odds ratio (with 95% confidence interval).

## DISCUSSION

In this study, we aimed to understand how the severity of a SARS-CoV-2 infection prior to major elective inpatient surgery influences postoperative outcomes using the N3C Data Enclave—a harmonized electronic health data platform comprising over 70 institutions and more than 18,000,000 patients^10^. Our findings demonstrate that a prior history of COVID-19 is an independent risk factor for adverse surgical outcomes, and the risk remains elevated for 12 weeks after a SARS-CoV-2 infection; however, this is dependent on the severity of COVID-19. Patients with mild disease (not requiring hospitalization) did not have an elevated risk of adverse outcomes regardless of the timing of surgery. Those with moderate and severe disease had persistently elevated surgical risk that lasted 12+ weeks after an acute infection.

The results of this study are consistent with prior work examining surgical outcomes in patients with a history of SARS-CoV-2 infection conducted early in the pandemic. The COVIDSurg Collaborative, a multinational collaborative that aimed to explore the impact of COVID-19 in surgical patients and services, published the first population-level study assessing surgical outcomes following a SARS-CoV-2 infection. They found an increased risk of surgical mortality that lasted for 7 weeks following an acute infection^7^. Another large-scale study by Deng and colleagues confirmed these findings in a United States population and advocated for a wait time of 8 weeks from the time of a SARS-CoV-2 infection before proceeding with elective surgery^5^. In response to these data (and other similar findings from single institution series), expert guidelines suggested delaying elective surgery 7 weeks while weighing the time-sensitivity of the intervention^13^. These studies and guidelines were crucial given the novel situation perioperative providers were faced with early in the COVID-19 pandemic as an increasing number of patients with prior SARS-CoV-2 infection required elective surgery.

Still, several gaps remain in the evidence base for guiding the management of this patient population which are now addressed by this study. We re-demonstrate the risk for adverse 30-day composite postoperative outcomes remains elevated for 8 weeks after a SARS-CoV-2 infection; however, when stratified by severity, this risk only is present in those recovered from moderate and severe infections (anyone who required hospitalization). Those with mild disease were not at increased risk of composite adverse outcomes even when undergoing surgery within 4 weeks of an acute infection which is consistent with prior work^8^. We also provide a more detailed measurement of the association across a spectrum of specific postoperative complications. For example, patients with mild disease had no increased odds for all assessed adverse events, moderate disease showed elevated risk for all assessed complications, while those with severe disease had increased odds of all assessed complications with a magnitude of effect multiple-fold greater. This study also provides evidence that vaccination can reduce the risk of adverse postoperative events in patients with a prior history of COVID-19.

The data presented in this study include patients who underwent surgery prior to March 2023 and thus reflects the current environment as the pandemic shifts to an endemic phase. As a comparison, data collected for the COVIDSurg study was in October of 2020, and the Deng et al. study of patients in the United States included surgeries performed until June of 2021. Equipped with the updated data presented in this study, we propose the following guidance for managing patients prior to inpatient elective surgery. First, all patients who are planned to undergo surgery should be assessed for their history of COVID-19, including detailed information regarding the severity of their infection. Those with a prior history should be classified as having either asymptomatic/mild disease or moderate/severe disease. After this determination is made, patients with asymptomatic/mild disease should have their surgery delayed at least 5 days from the time of positive test to ensure they do not develop progressive symptoms that increase the severity of illness. If they remain asymptomatic/mild after this wait time, our data supports the safety of proceeding with elective surgery.

However, if the patient has moderate/severe disease or develops moderate/severe disease, then consideration should be made for delaying surgery while also considering the risk/benefit ratio of potential harms related to a prolonged delay in circumstances of time-sensitive surgical indications. Since we do not observe a return to baseline risk even beyond 12 weeks in patients with moderate/severe disease, no standard time for delay can be recommended. Instead, we advocate for early engagement with perioperative medicine teams and/or dedicated long COVID clinics to identify strategies for optimal risk reduction and provide objective assessment of physiologic recovery prior to proceeding with surgery. At our institution, all patients with a history of moderate/severe disease are recommended to be referred to preoperative clinic for assessment prior to procedure to assess for recovery and help collaboratively determine the optimal timing for surgery. Developing and applying machine learning/artificial intelligence tools using the N3C Data Enclave are active areas of work in our group to help provide more specific guidance for patients with a history of moderate or severe SARS-CoV-2 infection.

There are several limitations to our study that must be carefully considered when interpreting the results. The use of electronic health record data within N3C Data Enclave may introduce biases that are well-described in prior studies^14^. Methods specific to N3C can help manage several of these through rigorous data standardization, a highly engaged user community, and a centralized platform for handling all data-related tasks including extraction, processing, and analyses. Another source of bias is the selection of patients to undergo surgery after recovering from a prior COVID-19 infection. Detailed information on preoperative evaluation, indications, and rationale for proceeding with surgery prior to 7 weeks in patients with prior COVID- 19 is not available. Additionally, we were unable to directly measure the association between SARS-CoV-2 variants and outcomes—though, this likely is indirectly accounted for in analyses that include severity. Despite these potential shortcomings, leveraging the largest COVID-19-specific national data provides a unique opportunity to help guide the management of patients with previous SARS-CoV-2 infections undergoing elective surgery.

In conclusion, prior COVID-19 infection is a risk factor for adverse postoperative outcomes following major inpatient elective surgery. The magnitude and duration of this risk is dependent on the severity of COVID-19 infection and is reduced in vaccinated patients. As the pandemic transitions to an endemic phase, the results of this study emphasize a one-sized-fits-all approach to risk stratification in this patient population can lead to unnecessary surgical delays, and consensus guidelines should be updated to include consideration of COVID-19 disease severity and vaccination status.

## Data Availability

All data produced are available using the N3C Data Enclave (covid.cd2h.org/enclave).

## DISCLAIMER

The N3C Publication committee confirmed that this manuscript (MSID: 1004.67) is in accordance with N3C data use and attribution policies; however, the content is solely the responsibility of the authors and does not necessarily represent the official views of the NIH. Authorship was determined using ICMJE recommendations.

## ACKNOWLEDGEMENT

The analyses described in this publication were conducted under the DUR RP-A39709 and IRB #PRO00042159 with data or tools accessed through the NCATS N3C Data Enclave covid.cd2h.org/enclave and supported by CD2H - The National COVID Cohort Collaborative (N3C) IDeA CTR Collaboration 3U24TR002306-04S2 NCATS U24 TR002306. The project described was supported by the National Center for Advancing Translational Sciences, National Institutes of Health, Award Number UL1 TR001436 and 3UL1TR001436-08S3. Additional funding support provided by Advancing a Healthier Wisconsin Endowment. This research was possible because of the patients whose information is included within the data from participating organizations (covid.cd2h.org/dtas) and the organizations and scientists (covid.cd2h.org/duas) who have contributed to the on-going development of this community resource (https://doi.org/10.1093/jamia/ocaa196).

We gratefully acknowledge the following core contributors to N3C:

Adam B. Wilcox, Adam M. Lee, Alexis Graves, Alfred (Jerrod) Anzalone, Amin Manna, Amit Saha, Amy Olex, Andrea Zhou, Andrew E. Williams, Andrew Southerland, Andrew T. Girvin, Anita Walden, Anjali A. Sharathkumar, Benjamin Amor, Benjamin Bates, Brian Hendricks, Brijesh Patel, Caleb Alexander, Carolyn Bramante, Cavin Ward-Caviness, Charisse Madlock-Brown, Christine Suver, Christopher Chute, Christopher Dillon, Chunlei Wu, Clare Schmitt, Cliff Takemoto, Dan Housman, Davera Gabriel, David A. Eichmann, Diego Mazzotti, Don Brown, Eilis Boudreau, Elaine Hill, Elizabeth Zampino, Emily Carlson Marti, Emily R. Pfaff, Evan French, Farrukh M Koraishy, Federico Mariona, Fred Prior, George Sokos, Greg Martin, Harold Lehmann, Heidi Spratt, Hemalkumar Mehta, Hongfang Liu, Hythem Sidky, J.W. Awori Hayanga, Jami Pincavitch, Jaylyn Clark, Jeremy Richard Harper, Jessica Islam, Jin Ge, Joel Gagnier, Joel H. Saltz, Joel Saltz, Johanna Loomba, John Buse, Jomol Mathew, Joni L. Rutter, Julie A. McMurry, Justin Guinney, Justin Starren, Karen Crowley, Katie Rebecca Bradwell, Kellie M. Walters, Ken Wilkins, Kenneth R. Gersing, Kenrick Dwain Cato, Kimberly Murray, Kristin Kostka, Lavance Northington, Lee Allan Pyles, Leonie Misquitta, Lesley Cottrell, Lili Portilla, Mariam Deacy, Mark M. Bissell, Marshall Clark, Mary Emmett, Mary Morrison Saltz, Matvey B. Palchuk, Melissa A. Haendel, Meredith Adams, Meredith Temple-O’Connor, Michael G. Kurilla, Michele Morris, Nabeel Qureshi, Nasia Safdar, Nicole Garbarini, Noha Sharafeldin, Ofer Sadan, Patricia A. Francis, Penny Wung Burgoon, Peter Robinson, Philip R.O. Payne, Rafael Fuentes, Randeep Jawa, Rebecca Erwin-Cohen, Rena Patel, Richard A. Moffitt, Richard L. Zhu, Rishi Kamaleswaran, Robert Hurley, Robert T. Miller, Saiju Pyarajan, Sam G. Michael, Samuel Bozzette, Sandeep Mallipattu, Satyanarayana Vedula, Scott Chapman, Shawn T. O’Neil, Soko Setoguchi, Stephanie S. Hong, Steve Johnson, Tellen D. Bennett, Tiffany Callahan, Umit Topaloglu, Usman Sheikh, Valery Gordon, Vignesh Subbian, Warren A. Kibbe, Wenndy Hernandez, Will Beasley, Will Cooper, William Hillegass, Xiaohan Tanner Zhang. Details of contributions available at covid.cd2h.org/core-contributors

The following institutions whose data is released or pending:

Available: Advocate Health Care Network — UL1TR002389: The Institute for Translational Medicine (ITM) • Boston University Medical Campus — UL1TR001430: Boston University Clinical and Translational Science Institute • Brown University — U54GM115677: Advance Clinical Translational Research (Advance-CTR) • Carilion Clinic — UL1TR003015: iTHRIV Integrated Translational health Research Institute of Virginia • Charleston Area Medical Center — U54GM104942: West Virginia Clinical and Translational Science Institute (WVCTSI) • Children’s Hospital Colorado — UL1TR002535: Colorado Clinical and Translational Sciences Institute • Columbia University Irving Medical Center — UL1TR001873: Irving Institute for Clinical and Translational Research • Duke University — UL1TR002553: Duke Clinical and Translational Science Institute • George Washington Children’s Research Institute — UL1TR001876: Clinical and Translational Science Institute at Children’s National (CTSA-CN) • George Washington University — UL1TR001876: Clinical and Translational Science Institute at Children’s National (CTSA-CN) • Indiana University School of Medicine — UL1TR002529: Indiana Clinical and Translational Science Institute • Johns Hopkins University — UL1TR003098: Johns Hopkins Institute for Clinical and Translational Research • Loyola Medicine — Loyola University Medical Center • Loyola University Medical Center — UL1TR002389: The Institute for Translational Medicine (ITM) • Maine Medical Center — U54GM115516: Northern New England Clinical & Translational Research (NNE-CTR) Network • Massachusetts General Brigham — UL1TR002541: Harvard Catalyst • Mayo Clinic Rochester — UL1TR002377: Mayo Clinic Center for Clinical and Translational Science (CCaTS) • Medical University of South Carolina — UL1TR001450: South Carolina Clinical & Translational Research Institute (SCTR) • Montefiore Medical Center — UL1TR002556: Institute for Clinical and Translational Research at Einstein and Montefiore • Nemours — U54GM104941: Delaware CTR ACCEL Program • NorthShore University HealthSystem — UL1TR002389: The Institute for Translational Medicine (ITM) • Northwestern University at Chicago — UL1TR001422: Northwestern University Clinical and Translational Science Institute (NUCATS) • OCHIN — INV- 018455: Bill and Melinda Gates Foundation grant to Sage Bionetworks • Oregon Health & Science University — UL1TR002369: Oregon Clinical and Translational Research Institute • Penn State Health Milton S. Hershey Medical Center — UL1TR002014: Penn State Clinical and Translational Science Institute • Rush University Medical Center — UL1TR002389: The Institute for Translational Medicine (ITM) • Rutgers, The State University of New Jersey — UL1TR003017: New Jersey Alliance for Clinical and Translational Science • Stony Brook University — U24TR002306 • The Ohio State University — UL1TR002733: Center for Clinical and Translational Science • The State University of New York at Buffalo — UL1TR001412: Clinical and Translational Science Institute • The University of Chicago — UL1TR002389: The Institute for Translational Medicine (ITM) • The University of Iowa — UL1TR002537: Institute for Clinical and Translational Science • The University of Miami Leonard M. Miller School of Medicine — UL1TR002736: University of Miami Clinical and Translational Science Institute • The University of Michigan at Ann Arbor — UL1TR002240: Michigan Institute for Clinical and Health Research • The University of Texas Health Science Center at Houston — UL1TR003167: Center for Clinical and Translational Sciences (CCTS) • The University of Texas Medical Branch at Galveston — UL1TR001439: The Institute for Translational Sciences • The University of Utah — UL1TR002538: Uhealth Center for Clinical and Translational Science • Tufts Medical Center — UL1TR002544: Tufts Clinical and Translational Science Institute • Tulane University — UL1TR003096: Center for Clinical and Translational Science • University Medical Center New Orleans — U54GM104940: Louisiana Clinical and Translational Science (LA CaTS) Center • University of Alabama at Birmingham — UL1TR003096: Center for Clinical and Translational Science • University of Arkansas for Medical Sciences — UL1TR003107: UAMS Translational Research Institute • University of Cincinnati — UL1TR001425: Center for Clinical and Translational Science and Training • University of Colorado Denver, Anschutz Medical Campus — UL1TR002535: Colorado Clinical and Translational Sciences Institute • University of Illinois at Chicago — UL1TR002003: UIC Center for Clinical and Translational Science • University of Kansas Medical Center — UL1TR002366: Frontiers: University of Kansas Clinical and Translational Science Institute • University of Kentucky — UL1TR001998: UK Center for Clinical and Translational Science • University of Massachusetts Medical School Worcester — UL1TR001453: The UMass Center for Clinical and Translational Science (UMCCTS) • University of Minnesota — UL1TR002494: Clinical and Translational Science Institute • University of Mississippi Medical Center — U54GM115428: Mississippi Center for Clinical and Translational Research (CCTR) • University of Nebraska Medical Center — U54GM115458: Great Plains IDeA-Clinical & Translational Research • University of North Carolina at Chapel Hill — UL1TR002489: North Carolina Translational and Clinical Science Institute • University of Oklahoma Health Sciences Center — U54GM104938: Oklahoma Clinical and Translational Science Institute (OCTSI) • University of Rochester — UL1TR002001: UR Clinical & Translational Science Institute • University of Southern California — UL1TR001855: The Southern California Clinical and Translational Science Institute (SC CTSI) • University of Vermont — U54GM115516: Northern New England Clinical & Translational Research (NNE- CTR) Network • University of Virginia — UL1TR003015: iTHRIV Integrated Translational health Research Institute of Virginia • University of Washington — UL1TR002319: Institute of Translational Health Sciences • University of Wisconsin-Madison — UL1TR002373: UW Institute for Clinical and Translational Research • Vanderbilt University Medical Center — UL1TR002243: Vanderbilt Institute for Clinical and Translational Research • Virginia Commonwealth University — UL1TR002649: C. Kenneth and Dianne Wright Center for Clinical and Translational Research • Wake Forest University Health Sciences — UL1TR001420: Wake Forest Clinical and Translational Science Institute • Washington University in St. Louis — UL1TR002345: Institute of Clinical and Translational Sciences • Weill Medical College of Cornell University — UL1TR002384: Weill Cornell Medicine Clinical and Translational Science Center • West Virginia University — U54GM104942: West Virginia Clinical and Translational Science Institute (WVCTSI) Submitted: Icahn School of Medicine at Mount Sinai — UL1TR001433: ConduITS Institute for Translational Sciences • The University of Texas Health Science Center at Tyler — UL1TR003167: Center for Clinical and Translational Sciences (CCTS) • University of California, Davis — UL1TR001860: UCDavis Health Clinical and Translational Science Center • University of California, Irvine — UL1TR001414: The UC Irvine Institute for Clinical and Translational Science (ICTS) • University of California, Los Angeles — UL1TR001881: UCLA Clinical Translational Science Institute • University of California, San Diego — UL1TR001442: Altman Clinical and Translational Research Institute • University of California, San Francisco — UL1TR001872: UCSF Clinical and Translational Science Institute Pending: Arkansas Children’s Hospital — UL1TR003107: UAMS Translational Research Institute • Baylor College of Medicine — None (Voluntary) • Children’s Hospital of Philadelphia — UL1TR001878: Institute for Translational Medicine and Therapeutics • Cincinnati Children’s Hospital Medical Center — UL1TR001425: Center for Clinical and Translational Science and Training • Emory University — UL1TR002378: Georgia Clinical and Translational Science Alliance • HonorHealth — None (Voluntary) • Loyola University Chicago — UL1TR002389: The Institute for Translational Medicine (ITM) • Medical College of Wisconsin — UL1TR001436: Clinical and Translational Science Institute of Southeast Wisconsin • MedStar Health Research Institute — UL1TR001409: The Georgetown-Howard Universities Center for Clinical and Translational Science (GHUCCTS) • MetroHealth — None (Voluntary) • Montana State University — U54GM115371: American Indian/Alaska Native CTR • NYU Langone Medical Center — UL1TR001445: Langone Health’s Clinical and Translational Science Institute • Ochsner Medical Center — U54GM104940: Louisiana Clinical and Translational Science (LA CaTS) Center • Regenstrief Institute — UL1TR002529: Indiana Clinical and Translational Science Institute • Sanford Research — None (Voluntary) • Stanford University — UL1TR003142: Spectrum: The Stanford Center for Clinical and Translational Research and Education • The Rockefeller University — UL1TR001866: Center for Clinical and Translational Science • The Scripps Research Institute — UL1TR002550: Scripps Research Translational Institute • University of Florida — UL1TR001427: UF Clinical and Translational Science Institute • University of New Mexico Health Sciences Center — UL1TR001449: University of New Mexico Clinical and Translational Science Center • University of Texas Health Science Center at San Antonio — UL1TR002645: Institute for Integration of Medicine and Science • Yale New Haven Hospital — UL1TR001863: Yale Center for Clinical Investigation

## SUPPLEMENTARY DATA

**Supplemental Table 1:** Surgical Procedures and associated risk.

| **Procedure Name** | **Surgery N=387,030 (%)** | **SNOMED Code** |
| --- | --- | --- |
| Low Risk |  |  |
| **Hip Arthroplasty** | 5480 (1.4%) | 52734007 |
| **Knee Arthroplasty** | 6034 (1.6%) | 609588000 |
| **Laminectomy** | 44462 (11%) | 387731002 |
| **Mastectomy** | 75714 (20%) | 69031006 |
| **Prostatectomy** | 17059 (4.4%) | 90470006 |
| Medium Risk |  |  |
| **Aortic Aneurysm Repair** | 4881 (1.3%) | 233370007 |
| **Colectomy** | 46059 (12%) | 23968004 |
| **Craniectomy** | 29201 (7.5%) | 36910002 |
| **Gastrectomy** | 7903 (2.0%) | 53442002 |
| **Lung Excision** | 17353 (4.5%) | 119746007 |
| **Spinal Arthrodesis** | 78947 (20%) | 55705006 |
| High Risk |  |  |
| **Coronary Artery Bypass Graft** | 37492 (9.7%) | 232717009 |
| **Esophagectomy** | 1538 (0.4%) | 45900003 |
| **Hepatectomy** | 8495 (2.2%) | 107963000 |
| **Pancreatectomy** | 6412 (1.7%) | 33149006 |

**Supplemental Table 2:** Patient comorbidities.

| Preoperative Comorbidities | SNOMED Codes |
| --- | --- |
| Chronic Kidney Disease | 709044004 |
| Chronic Obstructive Pulmonary Disease | 13645005 |
| Coronary Artery Disease | 53741008 |
| Depression | 370143000 |
| Diabetes | 73211009 |
| Gastroesophageal Reflux Disease | 235595009 |
| Heart Failure | 84114007 |
| Hypertension | 59621000 |
| Liver Disease | 328383001 |
| Obesity | 414916001 |

**Supplemental Table 3:**
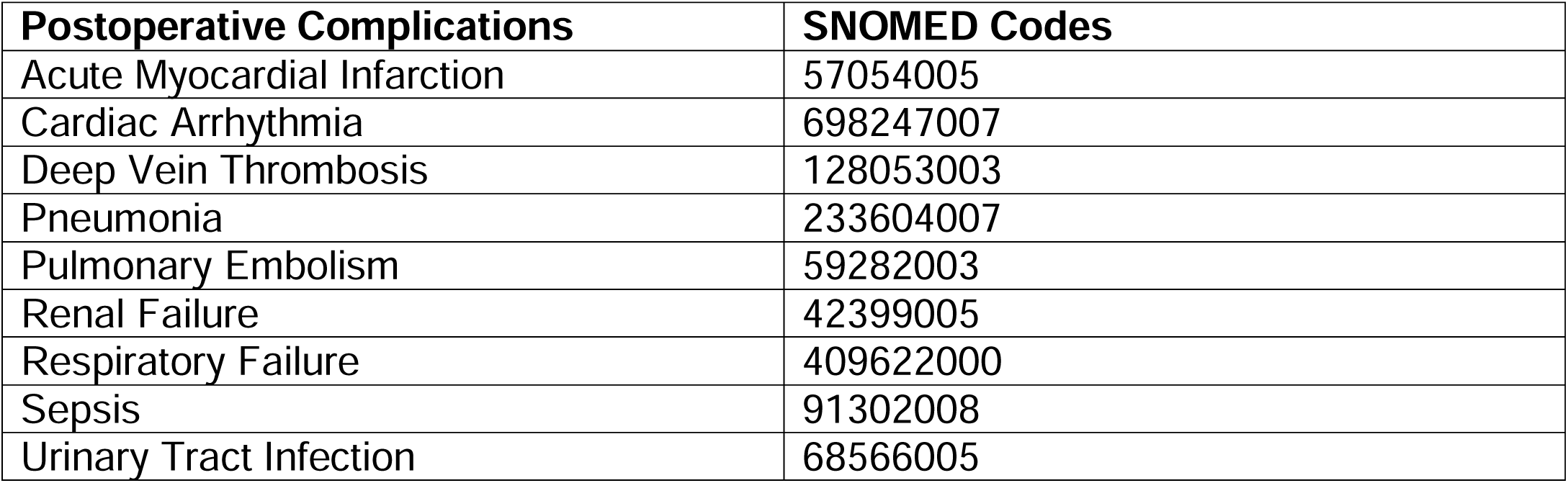
Postoperative Complications.

| Postoperative Complications | SNOMED Codes |
| --- | --- |
| Acute Myocardial Infarction | 57054005 |
| Cardiac Arrhythmia | 698247007 |
| Deep Vein Thrombosis | 128053003 |
| Pneumonia | 233604007 |
| Pulmonary Embolism | 59282003 |
| Renal Failure | 42399005 |
| Respiratory Failure | 409622000 |
| Sepsis | 91302008 |
| Urinary Tract Infection | 68566005 |

**Supplemental Table 4.** Comparison of 30-day postoperative adverse events based on surgical timing relative to COVID-19.

| Outcomes | No History of COVID-19 (N = 352,947) | 0–4 Weeks Post- COVID-19 (N = 7,425) | 4–8 Weeks Post- COVID-19 (N = 3,936) | 8–12 Weeks Post COVID-19 (N = 2,604) | 12+ Weeks Post COVID-19 (N = 23,389) | P value |
| --- | --- | --- | --- | --- | --- | --- |
| Mortality | 4791 (1.4%) | 246 (3.3%) | 83 (2.1%) | 47 (1.8%) | 290 (1.2%) | <0.001 |
| LOS | 2 (0, 6) | 4 (1, 8) | 2 (0, 6) | 3 (0, 6) | 2 (0, 5) | <0.001 |
| Hospital Readmission | 38175 (11%) | 848 (11%) | 458 (12%) | 318 (12%) | 2622 (11%) | 0.057 |
| Any Non-Fatal Complication | 73555 (21%) | 1914 (26%) | 890 (23%) | 625 (24%) | 4828 (21%) | <0.001 |
| Acute Myocardial Infarction | 9089 (2.6%) | 271 (3.6%) | 100 (2.5%) | 57 (2.2%) | 592 (2.5%) | <0.001 |
| Cardiac Arrhythmia | 34326 (9.8%) | 803 (11%) | 409 (10%) | 286 (11%) | 2254 (9.6%) | 0.006 |
| Deep Vein Thrombosis | 4648 (1.3%) | 142 (1.9%) | 57 (1.4%) | 46 (1.8%) | 287 (1.2%) | <0.001 |
| Pneumonia | 9623 (2.8%) | 348 (4.7%) | 131 (3.3%) | 95 (3.6%) | 519 (2.2%) | <0.001 |
| Pulmonary Embolism | 4296 (1.2%) | 137 (1.8%) | 58 (1.5%) | 35 (1.3%) | 289 (1.2%) | <0.001 |
| Renal Failure | 22488 (6.4%) | 654 (8.8%) | 288 (7.3%) | 204 (7.8%) | 1446 (6.2%) | <0.001 |
| Respiratory Failure | 18862 (5.4%) | 655 (8.8%) | 197 (5.0%) | 164 (6.3%) | 1001 (4.3%) | <0.001 |
| Sepsis | 10244 (2.9%) | 346 (4.7%) | 134 (3.4%) | 95 (3.6%) | 613 (2.6%) | <0.001 |
| Urinary Tract Infection | 9662 (2.8%) | 208 (2.8%) | 110 (2.8%) | 88 (3.4%) | 639 (2.7%) | 0.43 |

**Supplemental Table 5.** Multivariable regression assessing the impact of timing of infection relative to surgery on risk on composite and specific 30-day postoperative adverse outcomes. All models adjusted for age, sex, race and ethnicity, smoking status, and relative risk of surgery.

|  | **0–4 weeks**<br>**(N = 7,425)** |  | **4–8 weeks**<br>**(N = 3,936)** |  | **8–12 weeks**<br>**(N = 2,604)** |  | **12+ Weeks**<br>**(N = 23,389)** |  |
| --- | --- | --- | --- | --- | --- | --- | --- | --- |
| **Composite 30-day postoperative outcome** | 1.27 | [1.20-1.35] | 1.14 | [1.05-1.24] | 1.16 | [1.05-1.28] | 1.01 | [0.98-1.05] |
| **Mortality** | 2.34 | [2.05-2.67] | 1.57 | [1.25-1.95] | 1.22 | [0.90-1.62] | 0.92 | [0.81-1.03] |
| **Composite Non-fatal Complications** | 1.22 | [1.15-1.29] | 1.10 | [1.01-1.20] | 1.14 | [1.03-1.26] | 1.01 | [0.98-1.05] |
| **Hospital Readmission** | 0.99 | [0.92-1.07] | 1.02 | [0.92-1.13] | 1.06 | [0.94-1.19] | 1.01 | [0.97-1.05] |
| **Myocardial Infarction** | 1.13 | [0.99-1.29] | 0.93 | [0.75-1.14] | 0.73 | [0.55-0.96] | 1.06 | [0.96-1.15] |
| **Cardiac Arrhythmia** | 1.04 | [0.96-1.13] | 1.11 | [1.00-1.24] | 1.15 | [1.00-1.31] | 1.09 | [1.04-1.15] |
| **Deep Vein Thrombosis** | 1.33 | [1.12-1.57] | 1.04 | [0.79-1.34] | 1.21 | [0.89-1.61] | 0.88 | [0.78-0.99] |
| **Pneumonia** | 1.60 | [1.43-1.79] | 1.21 | [1.01-1.44] | 1.24 | [1.00-1.52] | 0.81 | [0.74-0.88] |
| **Pulmonary Embolism** | 1.44 | [1.20-1.70] | 1.17 | [0.89-1.50] | 1.01 | [0.71-1.39] | 0.96 | [0.85-1.08] |
| **Renal Failure** | 1.28 | [1.17-1.39] | 1.13 | [0.99-1.27] | 1.12 | [0.96-1.30] | 0.94 | [0.89-1.00] |
| **Respiratory Failure** | 1.53 | [1.40-1.66] | 0.90 | [0.77-1.04] | 1.09 | [0.92-1.28] | 0.80 | [0.74-0.85] |
| **Sepsis** | 1.52 | [1.36-1.69] | 1.14 | [0.95-1.36] | 1.15 | [0.92-1.40] | 0.87 | [0.80-0.94] |
| **Urinary Tract Infection** | 1.01 | [0.87-1.15] | 1.02 | [0.84-1.23] | 1.17 | [0.94-1.44] | 0.97 | [0.90-1.05] |

**Supplemental Table 6.** Comparison of specific 30-day postoperative adverse events based on COVID-19 Severity.

| Postoperative Complications | No History of COVID-19 (N = 351,857) | Mild Outpatient WHO Severity 1–3 (N = 31,677) | Moderate Hospitalized without invasive ventilation WHO Severity 4–6 (N= 5,009) | Severe Hospitalized with invasive ventilation or ECMO WHO Severity 7–9 (N= 655) | P value |
| --- | --- | --- | --- | --- | --- |
| Mortality | 4791 (1.4%) | 403 (1.3%) | 169 (3.4%) | 94 (14%) | <0.001 |
| LOS | 2 (0, 6) | 2 (0, 5) | 4 (1, 9) | 12 (4, 29) | <0.001 |
| Hospital Readmission | 38175 (11%) | 3293 (10%) | 832 (17%) | 121 (18%) | <0.001 |
| Any Non-Fatal Complication | 73555 (21%) | 6110 (19%) | 1746 (35%) | 396 (60%) | <0.001 |
| Acute Myocardial Infarction | 9089 (2.6%) | 723 (2.3%) | 243 (4.9%) | 53 (8.1%) | <0.001 |
| Cardiac Arrhythmia | 34326 (9.8%) | 2856 (9.0%) | 751 (15%) | 142 (22%) | <0.001 |
| Deep Vein Thrombosis | 4648 (1.3%) | 364 (1.1%) | 126 (2.5%) | 42 (6.4%) | <0.001 |
| Pneumonia | 9623 (2.8%) | 704 (2.2%) | 249 (5.0%) | 138 (21%) | <0.001 |
| Pulmonary Embolism | 4296 (1.2%) | 388 (1.2%) | 99 (2.0%) | 32 (4.9%) | <0.001 |
| Renal Failure | 22488 (6.4%) | 1822 (5.8%) | 588 (12%) | 180 (27%) | <0.001 |
| Respiratory Failure | 18862 (5.4%) | 1338 (4.2%) | 448 (8.9%) | 228 (35%) | <0.001 |
| Sepsis | 10244 (2.9%) | 764 (2.4%) | 271 (5.4%) | 151 (23%) | <0.001 |
| Urinary Tract Infection | 9662 (2.8%) | 791 (2.5%) | 214 (4.3%) | 39 (6.0%) | <0.001 |

**Supplemental Table 7.** Multivariable regression assessing the impact of COVID-19 disease severity on composite and specific 30-day postoperative adverse outcomes. All models adjusted for age, sex, race and ethnicity, smoking status, and relative risk of surgery.

|  | Mild<br>(N = 31,677) | Moderate<br>(N = 5,009) | Severe<br>(N = 655) |
| --- | --- | --- | --- |
| **Composite 30-day postoperative outcome** | 0.96 [0.93-1.00] | 1.52 [1.43-1.62] | 4.76 [4.01-5.68] |
| **Mortality** | 1.00 [0.90-1.11] | 1.77 [1.51-2.07] | 7.49 [5.93-9.35] |
| **Composite Non-fatal Complications** | 0.96 [0.93-0.99] | 1.47 [1.38-1.57] | 4.08 [3.44-4.84] |
| **Hospital Readmission** | 0.96 [0.92-1.00] | 1.26 [1.17-1.36] | 1.20 [0.98-1.46] |
| **Myocardial Infarction** | 0.94 [0.86-1.02] | 1.31 [1.13-1.50] | 2.02 [1.46-2.73] |
| **Cardiac Arrhythmia** | 1.03 [0.98-1.07] | 1.27 [1.17-1.38] | 2.02 [1.64-2.47] |
| **Deep Vein Thrombosis** | 0.87 [0.78-0.97] | 1.37 [1.13-1.63] | 3.01 [2.16-4.08] |
| **Pneumonia** | 0.85 [0.79-0.92] | 1.30 [1.14-1.48] | 5.53 [4.53-6.70] |
| **Pulmonary Embolism** | 1.01 [0.91-1.12] | 1.19 [0.96-1.45] | 2.56 [1.75-3.60] |
| **Renal Failure** | 0.93 [0.88-0.98] | 1.28 [1.17-1.40] | 3.24 [2.69-3.89] |
| **Respiratory Failure** | 0.82 [0.77-0.87] | 1.20 [1.08-1.32] | 5.71 [4.81-6.76] |
| **Sepsis** | 0.85 [0.78-0.91] | 1.33 [1.17-1.51] | 5.91 [4.88-7.12] |
| **Urinary Tract Infection** | 0.93 [0.86-1.00] | 1.25 [1.08-1.43] | 1.72 [1.22-2.36] |

**Supplemental Table 8.** Comparison of specific 30-day postoperative adverse events based on vaccination in patients with COVID-19.

| **Postoperative Complications** | **Non-Fully Vaccinated**<br>**(N = 31,973),**<br>**No. (%)** | **Fully Vaccinated**<br>**(N = 5,381),**<br>**No. (%)** | **P value** |
| --- | --- | --- | --- |
| **Mortality** | 585 (1.8%) | 81 (1.5%) | 0.11 |
| **LOS** | 2 (0, 6) | 3 (0, 6) | 0.014 |
| **Hospital Readmission** | 3644 (11%) | 604 (11%) | 0.73 |
| **Any Non-Fatal Complication** | 7072 (22%) | 1185 (22%) | 0.89 |
| **Acute Myocardial Infarction** | 892 (2.8%) | 128 (2.4%) | 0.10 |
| **Cardiac Arrhythmia** | 3172 (9.9%) | 580 (11%) | 0.056 |
| **Deep Vein Thrombosis** | 465 (1.5%) | 67 (1.2%) | 0.26 |
| **Pneumonia** | 970 (3.0%) | 123 (2.3%) | 0.003 |
| **Pulmonary Embolism** | 434 (1.4%) | 85 (1.6%) | 0.22 |
| **Renal Failure** | 2245 (7.0%) | 347 (6.4%) | 0.13 |
| **Respiratory Failure** | 1792 (5.6%) | 225 (4.2%) | <0.001 |
| **Sepsis** | 1042 (3.3%) | 146 (2.7%) | 0.039 |
| **Urinary Tract Infection** | 873 (2.7%) | 172 (3.2%) | 0.061 |

